# Reduced exercise capacity, chronotropic incompetence, and early systemic inflammation in cardiopulmonary phenotype Long COVID

**DOI:** 10.1101/2022.05.17.22275235

**Authors:** Matthew S. Durstenfeld, Michael J. Peluso, Punita Kaveti, Christopher Hill, Danny Li, Erica Sander, Shreya Swaminathan, Victor M. Arechiga, Scott Lu, Sarah A Goldberg, Rebecca Hoh, Ahmed Chenna, Brandon C. Yee, John W. Winslow, Christos J. Petropoulos, J. Daniel Kelly, David V. Glidden, Timothy J. Henrich, Jeffrey N. Martin, Yoo Jin Lee, Mandar A. Aras, Carlin S. Long, Donald J. Grandis, Steven G. Deeks, Priscilla Y. Hsue

## Abstract

**BACKGROUND:** Mechanisms underlying persistent cardiopulmonary symptoms following SARS-CoV-2 infection (post-acute sequelae of COVID-19 “PASC” or “Long COVID”) remain unclear. This study sought to elucidate mechanisms of cardiopulmonary symptoms and reduced exercise capacity using advanced cardiac testing.

**METHODS:** We performed cardiopulmonary exercise testing (CPET), cardiac magnetic resonance imaging (CMR) and ambulatory rhythm monitoring among adults > 1 year after confirmed SARS-CoV-2 infection in Long-Term Impact of Infection with Novel Coronavirus cohort (LIINC; substudy of NCT04362150). Adults who completed a research echocardiogram (at a median 6 months after SARS-CoV-2 infection) without evidence of heart failure or pulmonary hypertension were asked to complete additional cardiopulmonary testing approximately 1 year later. Although participants were recruited as a prospective cohort, to account for selection bias, the primary analyses were as a case-control study comparing those with and without persistent cardiopulmonary symptoms. We also correlated findings with previously measured biomarkers. We used logistic regression and linear regression models to adjust for potential confounders including age, sex, body mass index, time since SARS-CoV-2 infection, and hospitalization for acute SARS-CoV-2 infection, with sensitivity analyses adjusting for medical history.

**RESULTS:** Sixty participants (unselected for symptoms, median age 53, 42% female, 87% non-hospitalized) were studied at median 17.6 months following SARS-CoV-2 infection. On maximal CPET, 18/37 (49%) with symptoms had reduced exercise capacity (peak VO_2_<85% predicted) compared to 3/19 (16%) without symptoms (p=0.02). The adjusted peak VO_2_ was 5.2 ml/kg/min (95%CI 2.1-8.3; p=0.001) or 16.9% lower actual compared to predicted (95%CI 4.3- 29.6; p=0.02) among those with symptoms compared to those without symptoms. Chronotropic incompetence was present among 12/21 (57%) with reduced VO_2_ including 11/37 (30%) with symptoms and 1/19 (5%) without (p=0.04). Inflammatory markers (hsCRP, IL-6, TNF-α) and SARS-CoV-2 antibody levels measured early in PASC were negatively correlated with peak VO_2_ more than 1 year later. Late-gadolinium enhancement on CMR and arrhythmias on ambulatory monitoring were not present.

**CONCLUSIONS:** We found evidence of objectively reduced exercise capacity among those with cardiopulmonary symptoms more than 1 year following COVID-19, which was associated with elevated inflammatory markers early in PASC. Chronotropic incompetence may explain exercise intolerance among some with cardiopulmonary phenotype Long COVID.

**Graphical Abstract:** 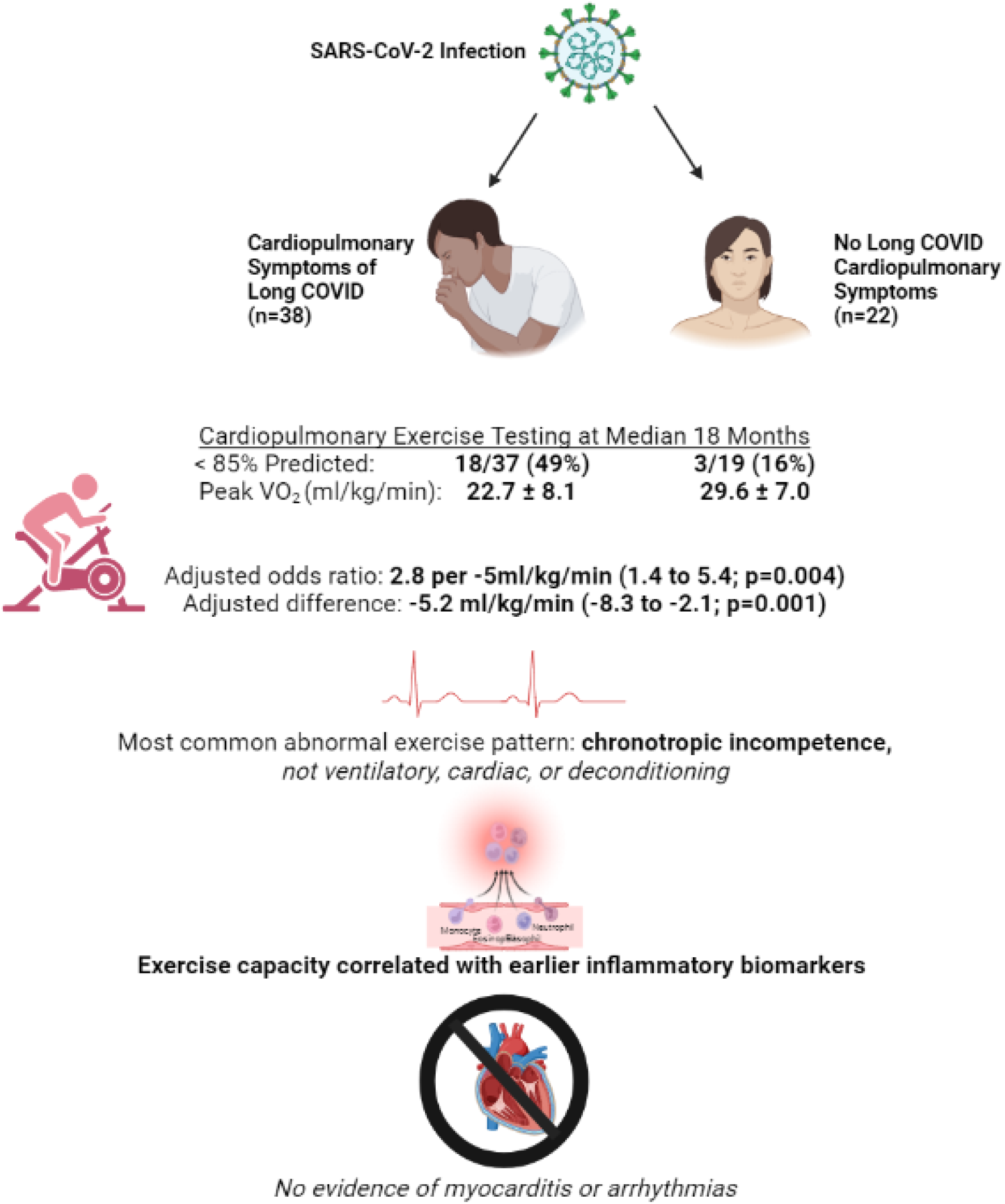

**Key Points:** Long COVID symptoms were associated with reduced exercise capacity on cardiopulmonary exercise testing more than 1 year after SARS-CoV-2 infection. The most common abnormal finding was chronotropic incompetence. Reduced exercise capacity was associated with early elevations in inflammatory markers.

## Background

Following acute SARS-CoV-2 infection, some individuals experience persistent symptoms of “Long COVID” (LC), a type of post-acute sequelae of COVID-19 (PASC) [1]. By 3-6 months after SARS-CoV-2 infection, cardiac function is generally normal on echocardiogram [2–4], suggesting that other techniques are needed to identify physiologic correlates of symptoms.

Cardiac magnetic resonance imaging (CMR) has revealed changes in parametric mapping and late gadolinium enhancement (LGE) suggestive of cardiac inflammation without consistent associations with symptoms or differences from controls [5–9]. Cardiopulmonary exercise testing (CPET) has demonstrated reduced exercise capacity 3-6 months after SARS-CoV-2 infection without consistent patterns of limitations [10], and with limited data beyond 1 year after infection.

We designed the Long-Term Impact of Infection with Novel Coronavirus (LIINC) study (NCT 04362150) to evaluate physical and mental health following SARS-COV-2 infection by including individuals representing the spectrum of acute illness and post-acute-recovery [11]. The purpose of this substudy was to elucidate mechanisms underlying cardiopulmonary symptoms >1 year following SARS-CoV-2 infection by comparing symptomatic and recovered individuals using CMR, CPET, and ambulatory rhythm monitoring, and correlating findings with blood-based markers.

## Methods

As previously reported, LIINC is a San Francisco-based post-COVID cohort [11]. After our initial echocardiogram-based study did not reveal cardiac mechanisms of symptoms [2], we amended our protocol to conduct a second visit about 1 year later among participants who completed an echocardiogram visit and were willing to undergo additional cardiopulmonary testing. In this subset, we performed cross-sectional cardiopulmonary testing including CPET, CMR, and ambulatory rhythm monitoring and correlated with already measured biomarkers from prior cohort visits (including at the time of the echocardiogram for troponin, NT-pro-BNP, and hsCRP).

### Participants

We invited LIINC participants with PCR-confirmed SARS-CoV-2 infection who completed an echocardiogram study visit to participate in additional cardiopulmonary testing irrespective of symptom status. Those with pregnancy (due to expected changes during pregnancy), cardiac disease (congenital heart disease, heart failure, myocardial infarction, coronary revascularization with percutaneous coronary intervention or coronary artery bypass graft surgery, or other heart surgery), pulmonary disease requiring home oxygen or lung surgery, and musculoskeletal or neurologic conditions that precluded participation in cycler ergometry were excluded.

Additionally, those with non-MRI compatible implants or claustrophobia were excluded from CMR; those with eGFR <30 ml/min/1.73m^2^ were excluded from gadolinium.

### Symptoms

Participants completed a structured interview about medical history, characteristics of acute infection, cardiopulmonary diagnoses, and symptoms within the previous two weeks. We defined cardiopulmonary symptoms as chest pain, dyspnea, or palpitations and symptoms as cardiopulmonary symptoms or fatigue in the 2 weeks preceding the study visit. Consistent with the WHO definition, all classified as symptomatic were >3 months after SARS-CoV-2 infection with new symptoms without alternative cardiopulmonary explanations [12].

### Blood-Based Markers

Participants had venous blood collected and processed for serum and plasma on the day of the echocardiogram. Samples were batch processed for measurement of high-sensitivity troponin I, N-terminal prohormone b-type natriuretic protein. A subset had antibodies and additional markers measured at two earlier time points (<90 days and 90-150 days after infection) including IL-6, IL-10, glial fibrillary acidic protein (GFAP), neurofilament light chain (NfL), monocyte chemoattractant protein-1 (MCP-1), interferon gamma (IFN -γ), and tumor necrosis factor (TNF), and SARS-CoV-2 receptor-binding domain (RBD) immunoglobulin G (IgG). These samples were assayed by Monogram Biosciences using the Quanterix Simoa® platform blinded with respect to patient and clinical information, and assay performance was consistent with manufacturers’ specifications [13].

### Cardiac Magnetic Resonance Imaging

Multiparametric, sequence-standardized, blinded (technician and reader) cardiac magnetic resonance imaging (CMR) was performed with a 3T system (Premier, General Electric), including assessment of LV and RV size and function, parametric mapping, and late gadolinium enhancement. Measurements were performed by a single reader at a dedicated workstation using Medis (Leiden, Netherlands) and AI-assisted Arterys (San Francisco, CA) under supervision of a senior cardiac imager, both blinded to clinical variables, and in accordance with Society for Cardiovascular Magnetic Resonance recommendations. The full protocol is described in Supplemental Methods.

### Cardiopulmonary Exercise Testing

Noninvasive CPETs were performed by an exercise physiologist and cardiology nurse practitioner blinded to participant data according to standard protocol using a metabolic cart (Medical Graphics Corporation Ultima CardiO_2_) and cycle ergometer (Lode Corival CPET) with continuous 12 lead ECG monitoring (GE CASE) and noninvasive blood pressure and pulse oximetry measurement. After rest ECG, BP and spirometry measurements were taken, participants exercised to symptom limited maximal exertional with work increased in 1-minute steps targeting a 10-minute test. We determined the work increase per 1 minute step based on the expected peak VO_2_ from the maximum voluntary ventilation for a goal 10 minute test, rounded to 5 Watts/min increments based on reported exercise (range 10-30 Watts/min) in accordance with guidelines [14]. The full protocol is described in Supplemental Methods.

We evaluated measured peak VO_2_ (in ml/kg/min), estimated percent predicted peak VO_2_ using the Wasserman equations [15], and classified peak VO_2_ <85% predicted as reduced. We defined chronotropic incompetence as peak VO_2_ <85% predicted, adjusted heart rate reserve (AHRR) <80% [(HR_peak_-HR_rest_)/(220-age-HR_rest_)], and no alternative explanation for exercise limitation [16]. CPETs were interpreted independently by two cardiologists with discrepancies resolved through consensus.

### Ambulatory Rhythm Monitoring

An ambulatory rhythm monitor (Carnation Ambulatory Monitor, BardyDx) was placed on participants’ chests. They were instructed wear it for 2 weeks, press the button for symptoms, and record a symptom diary. Monitors were processed according to BardyDx standard procedures, and reports were overread by a cardiologist.

### Statistical Analysis

To compare participants with and without symptoms, we used logistic regression to estimate adjusted odds ratios of parameters with symptoms and linear regression to estimate adjusted mean differences. Adjusted models included age, sex, time since SARS-CoV-2 infection, hospitalization, and body mass index. Non-normally distributed variables were log-transformed and findings are reported per doubling or 10-fold change. For biomarker data we report unadjusted Pearson’s rho correlation coefficients and adjusted linear regression models. For longitudinal data we used mixed effects models with random intercept per patient. We conducted sensitivity analyses considering other symptom definitions and additionally adjusting for potentially relevant medical history (hypertension, diabetes, asthma/COPD, and HIV) and echocardiographic (LV ejection fraction and LV diastolic function) and spirometry parameters (forced vital capacity, forced expiratory volume in 1 second, and maximal voluntary ventilation.

REDCap was used for data entry. Statistical analyses were performed using STATA version 17.1. P values <0.05 were considered statistically significant and were not adjusted for multiple testing. The first author (MSD) had full access to the data and takes responsibility for the integrity of the data and analysis. The study was approved by the UCSF Institutional Review Board (IRB 20-33000). All participants provided written informed consent prior to participation.

## Results

### Participant Characteristics

60 participants were included. Median age was 53 (IQR 41-59.5), 25 (42%) were female, and 8 (13%) were hospitalized during acute infection (Table 1). Median infection was in June 2020 (IQR March 2020-November 2020), so most participants were infected with the ancestral strain.

**Table 1:**
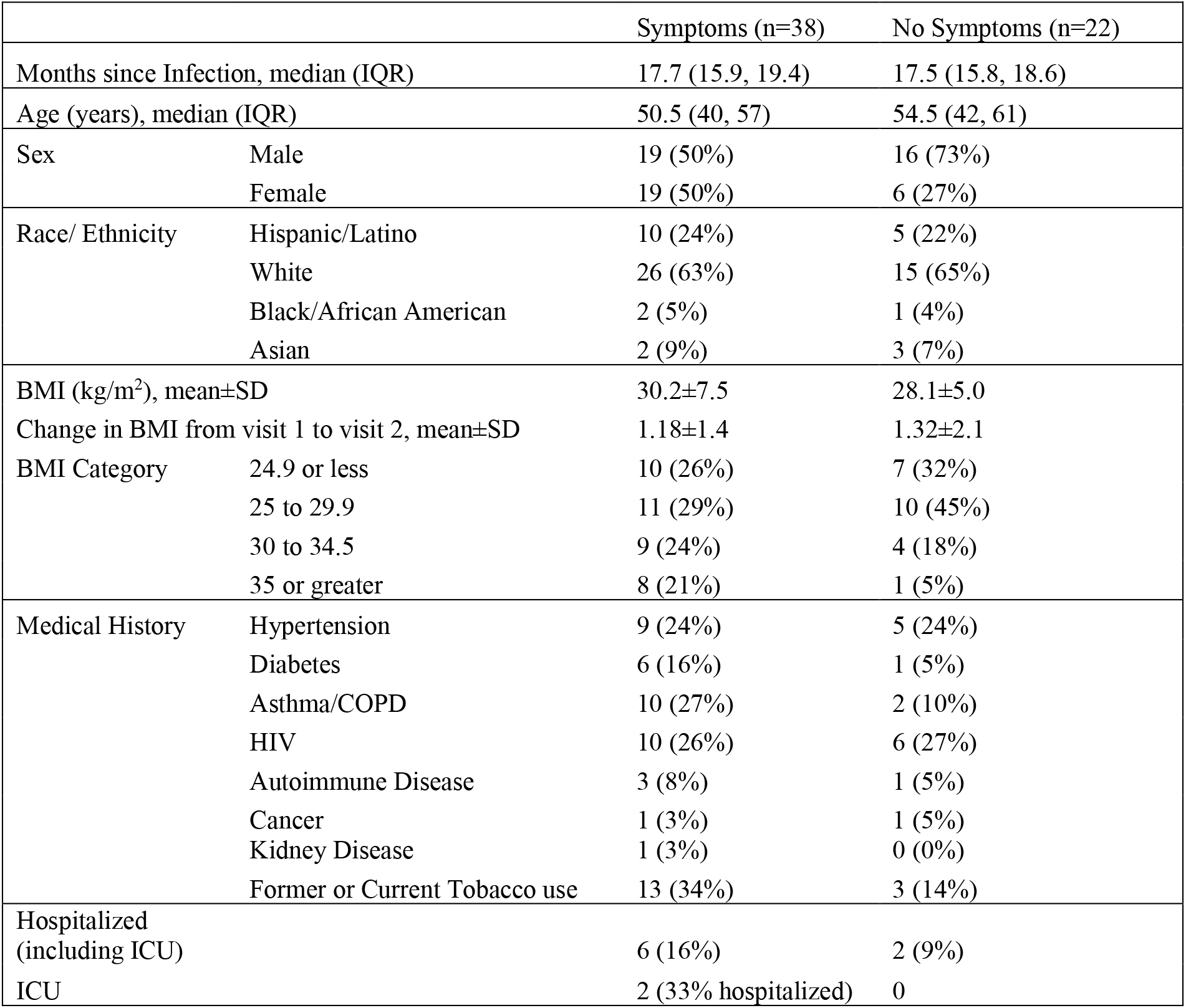
Baseline Characteristics (n=60) Table 1 **Legend:** Demographic information, past medical history, and severity of acute COVID-19 by hospitalization/ICU status of the participants who underwent advanced cardiopulmonary testing. Abbreviations: BMI=body mass index, ICU=intensive care unit, IQR=Interquartile Range

|  |  | Symptoms (n=38) | No Symptoms (n=22) |
| --- | --- | --- | --- |
| Months since Infection, median (IQR) |  | 17.7 (15.9, 19.4) | 17.5 (15.8, 18.6) |
| Age (years), median (IQR) |  | 50.5 (40, 57) | 54.5 (42, 61) |
| Sex | Male | 19 (50%) | 16 (73%) |
|  | Female | 19 (50%) | 6 (27%) |
| Race/ Ethnicity | Hispanic/Latino | 10 (24%) | 5 (22%) |
|  | White | 26 (63%) | 15 (65%) |
|  | Black/African American | 2 (5%) | 1 (4%) |
|  | Asian | 2 (9%) | 3 (7%) |
| BMI (kg/m<sup>2</sup>), mean±SD |  | 30.2±7.5 | 28.1±5.0 |
| Change in BMI from visit 1 to visit 2, mean±SD |  | 1.18±1.4 | 1.32±2.1 |
| BMI Category | 24.9 or less | 10 (26%) | 7 (32%) |
|  | 25 to 29.9 | 11 (29%) | 10 (45%) |
|  | 30 to 34.5 | 9 (24%) | 4 (18%) |
|  | 35 or greater | 8 (21%) | 1 (5%) |
| Medical History | Hypertension | 9 (24%) | 5 (24%) |
|  | Diabetes | 6 (16%) | 1 (5%) |
|  | Asthma/COPD | 10 (27%) | 2 (10%) |
|  | HIV | 10 (26%) | 6 (27%) |
|  | Autoimmune Disease | 3 (8%) | 1 (5%) |
|  | Cancer | 1 (3%) | 1 (5%) |
|  | Kidney Disease | 1 (3%) | 0 (0%) |
|  | Former or Current Tobacco use | 13 (34%) | 3 (14%) |
| Hospitalized (including ICU) |  | 6 (16%) | 2 (9%) |
| ICU |  | 2 (33% hospitalized) | 0 |

Four participants were vaccinated pre-infection (“breakthrough” infections), and 57 (95%) received at least one SARS-CoV-2 vaccine prior to advanced testing.

### Symptoms Persist at 18 Months

At visit 1 (median 6 months after infection; echocardiogram visit), 40/60 (67%) reported symptoms and 32/60 (53%) reported cardiopulmonary symptoms. At visit 2 (median 17.6 months; advanced cardiopulmonary testing visit), 38/60 (63%) reported symptoms and 31/60 (52%) reported cardiopulmonary symptoms. Trajectories of individual symptoms were similar (Supplemental Table 1). Self-reported reduced exercise capacity was highly associated with symptoms: 29/33 (88%) reporting reduced exercise capacity also reported other symptoms versus 9/27 (33%) reporting preserved or improved exercise capacity (OR 14.5, 95%CI 3.9-54.1; p<0.001).

### CPETs were Maximal Tests

Out of 60 participants who attended a CPET visit, 59 completed CPET at a median 17.6 months after infection (IQR 15.8-19.4); one participant was too hypertensive to undergo CPET. Out of 59 CPETs performed, one was excluded due to β-blocker use, two were excluded for submaximal tests with respiratory exchange ratio (RER) <1.05, leaving 56 CPETs for analysis. Three were stopped for hypertensive response (after reaching >100% predicted peak VO_2_); all others were symptom-limited maximal tests. No included participants were taking chronotropic medications or antianginals including β-blockers, non-dihydropyridine calcium channel blockers, ivabradine, or long-acting nitrates at the time of CPET.

### Exercise Capacity Lower than Predicted and Objectively Reduced Among those with Symptoms

Peak VO_2_ was <85% predicted among 18/37 (49%) with symptoms compared to 3/19 (16%) without symptoms (p=0.02). A 5 ml/kg/min decrease in peak VO_2_ was associated with 2.75 times higher odds of symptoms (95%CI 1.39-5.44; p=0.004). Those with symptoms completed less work despite higher perceived effort and similar respiratory exchange ratio (Table 2).

**Table 2:** Selected Cardiopulmonary Exercise Testing Parameters by Symptom Status (n=56) Table 2 **Legend:** We present both the odds ratios for the association between CPET parameters and symptoms estimated using logistic regression with adjustment for age, sex, time since COVID, hospitalization for acute COVID, BMI category and the estimated adjusted mean differences between those with and without symptoms using linear regression adjusting for the same covariates. Sensitivity analysis incorporating history of hypertension, diabetes, and lung disease had no substantive changes in effect sizes or confidence intervals. ^a^VE/VCO2 slope could not be determined for one participant without symptoms. ^b^Borg scale of perceived exertion was assessed every 2 minutes; these represent the last measurement prior to test stopping. **Bold text represents p<0.05.** Abbreviations: AT=Anaerobic threshold; bpm=beats per minute; FVC=Forced Vital Capacity; HR=heart rate; DBP=diastolic blood pressure; MVV=maximal voluntary ventilation; SBP=systolic blood pressure; VD/VT=Dead space ratio; VE = minute ventilation; VCO2=carbon dioxide production; pVO2=peak oxygen consumption (VO2); Vent=Ventilatory.

|  | Measure | Symptoms (n=37) | No Symptoms (n=19) | Adjusted OR (95%CI; p value) | Adjusted Difference (95%CI; p value) |
| --- | --- | --- | --- | --- | --- |
| **Exercise Capacity** | Peak VO<sub>2</sub>, ml/kg/min | 22.7 ± 8.1 | 29.6 ± 7.0 | 2.75 per -5 ml/kg/min (1.39-5.44; p=0.004) | -5.2 (-8.3to -2.1; p=0.001) |
|  | Peak VO<sub>2</sub>, % predicted | 92.0 ± 22.0 | 107.3 ± 22.0 | 1.22 per -5% (1.04-1.43; p=0.01) | -17 (-30 to -4.3; p=0.01) |
|  | Peak VO<sub>2</sub>, L/min | 1.9 ± 0.6 | 2.4 ± 0.7 | 1.18 per -0.1/min (1.03-1.37; p=0.02) | -0.41 (-0.73 to -0.09; p=0.01) |
|  | Proportion <85% predicted | 18 (49%) | 3 (16%) | 7.97 (1.56 to 40.8; p=0.01) | -- |
| **Ventila-tory** | Peak Respiratory Rate | 37.5 ± 8.7 | 40.8 ± 9.7 | 0.95 (0.88-1.02; p=0.17) | -3.8 (-10.0 to 2.4; p=0.23) |
|  | Breathing Reserve (MVV-V<sub>E</sub>max) | 44.5 (38.3, 59.0) | 32.8 (24.0, 44.8) | 1.04 (1.00-1.07; p=0.05) | 14.8 (0.7 to 29; p=0.04) |
|  | Vent. Efficiency (V<sub>E</sub>/VCO<sub>2</sub> slope)<sup>a</sup> | 27.5 ± 3.7 | 25.8 ± 3.7 | 1.18 (0.98-1.44; p=0.09) | 2.0 (-0.4 to 4.4; p=0.10) |
| **Peripheral** | VO<sub>2</sub> to Work slope | 9.9±3.0 | 10.9±3.5 | 0.94 (0.76-1.17; p=0.60) | -0.5 (-2.6 to 1.5; p=0.60) |
| **Cardiac** | VO<sub>2</sub> pulse, ml/beat | 13.1 ± 3.3 | 15.4 ± 4.5 | 0.85 (0.69-1.03; p=0.11) | -1.6 (-3.8 to 0.5; p=0.13) |
|  | SBP peak, mm Hg | 173.9 ± 31.7 | 189.2 ± 23.3 | 0.86 per 5 mm Hg (0.73-1.03; p=0.10) | -13.2 (-31.3 to 4.8; p=0.15) |
| **Heart Rate** | Rest, bpm | 78.3 ± 14.5 | 75.2 ± 10.4 | 1.01 (0.96-1.06; p=0.75) | 0.9 (-6.9 to 8.7; p=0.83) |
|  | Peak, bpm | 147.2 ± 25.7 | 154.1 ± 21.7 | 0.97 (0.94-1.01; p = 0.10) | -9.3 (-21 to 2; p=0.11) |
|  | Peak, % Age Predicted | 86.2 ± 11.9 | 94.1 ± 9.3 | 0.92 (0.85-0.99; p=0.02) | -8.5 (-15.2 to -1.8; p=0.02) |
|  | Adjusted HR Reserve Achieved, % | 73.6 ± 22.0 | 84.9 ± 21.2 | 0.97 (0.94-1.00; p=0.05) | -12.1 (-24.6 to 0.5; p=0.06) |
|  | HR Recovery at 1 min, bpm | 14.4 ± 7.4 | 14.0 ± 10.0 | 1.04 (0.95-1.14; p=0.39) | 2.0 (-2.9 to 6.9; p=0.42) |
| **Exertion** | Work (Watts) | 140.8 ± 60.6 | 196.2 ± 68.2 | 1.20 per -10 Watts (1.04 to 1.40; p=0.01) | -49.6 (-86.2 to -13.0; p=0.009) |
|  | Perceived Exertion, Borg Scale 6-20<sup>b</sup> | 16.2 ± 1.8 | 14.9 ± 2.2 | 1.64 (1.05-2.57; p=0.03) | 1.3 (0.1 to 2.6; p=0.03) |
|  | Peak Respiratory Exchange Ratio (VCO<sub>2</sub>/VO<sub>2</sub>) | 1.18 (1.12, 1.23) | 1.20 (1.12, 1.30) | 1.01 per -0.1 (0.48-2.12; p=0.98) | 0.03 (-0.09 to 0.02; p=0.23) |

Despite reduced exercise capacity among those with symptoms, most CPET parameters were not associated with symptoms (Table 2 and Supplemental Table 2 for additional parameters including rest spirometry & echocardiographic parameters).

As shown in Figure 1 and Table 2, peak VO_2_ was 22.7±8.1 and 29.6±7.0 ml/kg/min among those with and without symptoms, respectively, a difference of 6.9 ml/kg/min (95%CI 2.5-11.3; p=0.003) and 92% versus 107% percent predicted (difference 15% predicted, 95%CI; p=0.02).

**Figure 1.**
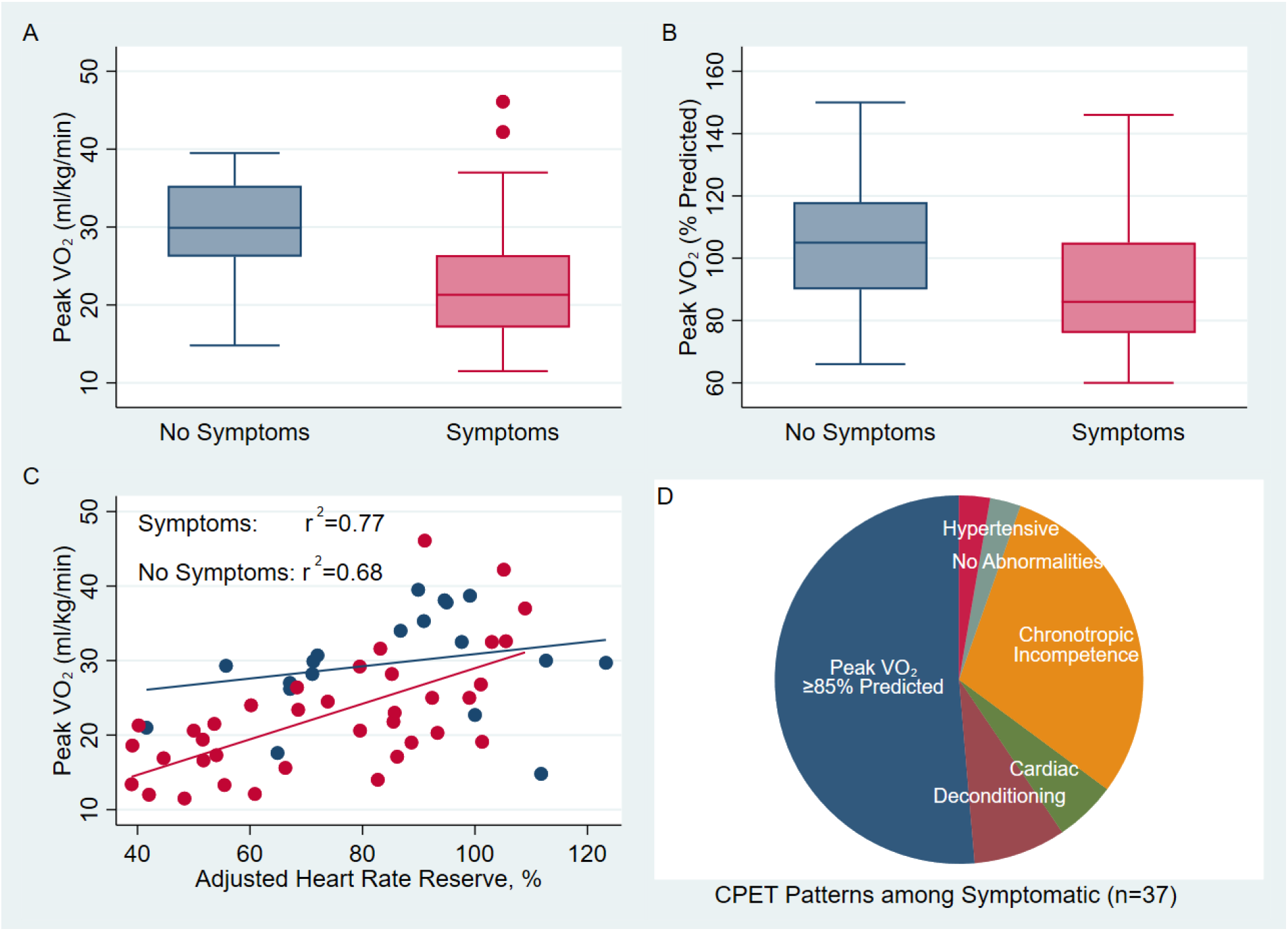
Exercise Capacity by Symptoms and HR Response to Exercise (n=56) Figure 1 **Legend:** Box and whisker plots of peak VO_2_ (ml/kg/min in Panel A and percent predicted in Panel B) among those without (blue) and with symptoms (pink) 17.6 months after SARS-CoV-2 infection (top). In Panel C peak VO_2_ in ml/kg/min is plotted by adjusted heart rate reserve (AHRR) to demonstrate the cluster of symptomatic individuals with low peak VO_2_ and chronotropic incompetence in the bottom left. Panel D demonstrates CPET patterns among those with Long COVID symptoms: half achieved greater than 85% predicted peak VO_2_, and chronotropic incompetence was the most common pattern among those with reduced exercise capacity.

The adjusted difference in peak VO_2_ was 5.2 ml/kg/min (95%CI 2.1-8.3; p=0.001), 0.4 L/min (95%CI 0.09 to 0.73; p=0.02), and 16.9% lower predicted (95%CI 4.3-29.6; p=0.02). Results were unchanged in sensitivity analysis adding diabetes and hypertension; the adjusted difference in peak VO_2_ was 4.5 ml/kg/min (95%CI 1.40-7.50; p=0.005). Similarly, results were unchanged in sensitivity analyses accounting for diabetes, hypertension, asthma/COPD, and HIV, resting spirometry values, and echocardiographic parameters (adjusted difference 3.9 ml/kg/min, 95%CI 0.6-7.3; p=0.02), but did vary based on symptom classification used (Supplemental Table 3).

### Classification of Reduced Exercise Capacity by Pattern of CPET Findings

Among 56 maximal CPETs, 21 (37%) had peak VO_2_<85% predicted; no participants had ventilatory limitation, 3 had cardiac limitation, and one had a hypertensive response. Four had findings most consistent with deconditioning/obesity, and one participant’s peak VO_2_ was 84% predicted with no other abnormalities (possibly deconditioning). Twelve (21% overall, 57% of those with reduced exercise capacity) had chronotropic incompetence. Among those with symptoms, 11/37 (30%) had chronotropic incompetence compared to 1/19 (5%) without symptoms (p=0.04).

Compared to those with peak VO_2_ ≥85% without an impaired chronotropic response, those with chronotropic incompetence had 49 bpm lower peak heart rate (119 bpm vs 170; 95%CI 40-60; p<0.0001; Figure 2). They completed 100 Watts less work (196 vs 96, 95%CI 49-152; p=0.0005) and had 12.2 ml/kg/min lower peak VO_2_ (95%CI 6.5-17.9; p=0.0001). Those with chronotropic incompetence also had reduced HR recovery at 1 minute (7.9 bpm lower, 95%CI 1.3-14.6; p=0.02). In absolute terms, those with chronotropic incompetence generated a mean peak oxygen consumption of 1.59 L/min compared to 2.35 L/min among those with normal exercise capacity (difference 0.76 L/min, 95%CI 0.23 to 1.28; p=0.007); a linear regression model with only rest and peak heart rate explains 54% of the difference in relative oxygen consumption (ml/kg/min) and 34% of the difference in absolute oxygen consumption (L/min).

**Figure 2.**
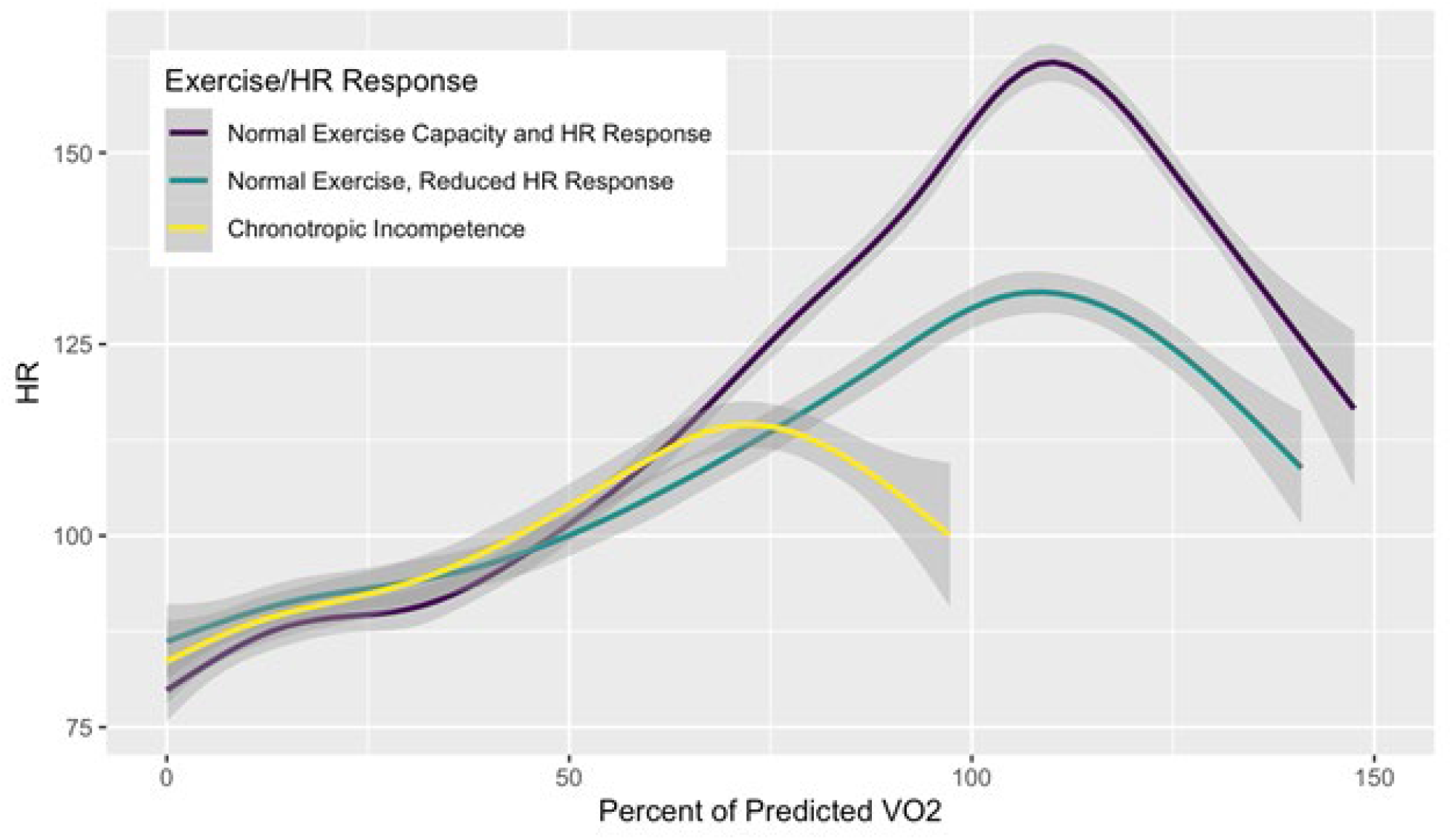
Heart Rate during Exercise by Chronotropic Response to Exercise. Figure 2 **Legend:** Mean heart rate is plotted as a function of exercise time normalized to percent of predicted peak VO_2_: in purple are those with normal exercise capacity (peak VO_2_ >85% predicted and normal heart rate response (n=16), in teal are those with normal exercise capacity (peak VO2>85%; n=8) and blunted heart rate response (AHRR<80%; n=8), and in yellow are those with chronotropic incompetence (n=9), as described in Supplemental Table 4.

### Normal Cardiac Structure and Function on CMR

Forty-three participants completed CMR, including two without gadolinium (one eGFR<30 and one due to inability to place an IV). CMR demonstrated normal LV and RV volumes and ejection fraction, and only RV volumes were associated with symptoms with smaller RV size associated with higher odds of symptoms (Table 3). No participants had LGE suggestive of myocardial scar, and native T1 and T2 parametric mapping values and ECV were not associated with symptoms. Some participants (10/43, 23%) had trace or small pericardial effusions with no difference by symptoms (p=0.59).

**Table 3.** Cardiac Magnetic Resonance Imaging Parameters (n=41) by Symptom Status. Table 3 **Legend:** CMR parameters by cardiopulmonary symptoms given as mean±SD or median (intraquartile range) for non-normally distributed variables. Logistic regression was used to estimated odds of having symptoms for a given change in each parameter adjusted for age, sex, BMI category, hospitalization, and time since infection. Only RV end diastolic and end systolic volume indices were associated with symptoms with larger RV size associated with lower odds of symptoms. **Bold text represents p<0.05.** Abbreviations: LVEDi=Left ventricular end diastolic volume indexed to body surface area; LVESi=Left ventricular end diastolic volume indexed to body surface area; LVEF=left ventricular ejection fraction; RVEDi=Right ventricular end diastolic volume indexed to body surface area; RVESi=Right ventricular end diastolic volume indexed to body surface area; RVEF=Right ventricular Ejection Fraction. LGE=Late Gadolinium Enhancement. ^a^LV stroke volumes are reported but there is a high correlation between LV and RV stroke volumes (Pearson’s r=0.96).

| Meaning | Parameter | Symptoms (n=25) | No Symptoms (n=18) | Adjusted OR (95%CI; p value) |
| --- | --- | --- | --- | --- |
| **Months since SARS-CoV-2 Infection, mean±SD** |  | 15.9±3.8 | 15.9±3.9 | -- |
| **Hematocrit, mean±SD** |  | 43.0 ± 3.5 | 44.5 ± 4.1 | 0.86 (0.67 to 1.09; p=0.21) |
| **Body Surface Area, m<sup>2</sup>** |  | 1.95 ± 0.22 | 1.90 ± 0.16 | 1.26 per 0.1 m<sup>2</sup> (0.74-2.17; p=0.38) |
| **Left Ventricular Size and Function** | LVEDi, ml/m<sup>2</sup> | 63.6 ± 13.9 | 69.3 ± 12.3 | 0.97 (0.91-1.03; p=0.27) |
|  | LVESi, ml/m<sup>2</sup> | 24.4 ± 6.9 | 25.3 ± 7.0 | 1.00 (0.89-1.12; p=0.97) |
|  | LVEF, % | 61.8 ± 5.9 | 63.3 ± 5.8 | 0.93 (0.82-1.05; p=0.26) |
|  | LV Mass Index, gm/m<sup>2</sup> | 47.6 ± 7.9 | 51.6 ± 7.6 | 0.98 (0.90 to 1.06; p=0.60) |
|  | Stroke Volume, ml<sup>a</sup> | 77.2 ± 18.1 | 84.2 ± 18.1 | 0.97 (0.93-1.01; p=0.20) |
| **Right Ventricular Size and Function** | **RVEDi, ml/m<sup>2</sup>** | **65.3 ± 13.5** | **75.6 ± 12.4** | **0.92 (0.86 to 0.99; p=0.02)** |
|  | **RVESi, ml/m<sup>2</sup>** | **27.1 ± 6.4** | **31.3 ± 7.6** | **0.86 (0.75=0.99; p=0.04)** |
|  | RVEF, % | 58.9 ± 5.0 | 58.4 ± 5.0 | 0.75 (0.86 to 1.14; p=0.93) |
| **Markers of Cardiac Inflammation** | T1 Native Mapping, ms | 1202 (1141, 1253) | 1219 (1153, 1248) | 1.00 (0.99-1.00; p=0.51) |
|  | Post-Contrast T1 Mapping Time, ms | 603 (507, 634) | 624 (577, 655) | 1.00 (0.99 to 1.01; p=0.64) |
|  | Extracellular Volume, % | 26.7 ± 6.3 | 24.2 ± 5.3 | 1.08 (0.92-1.26; p=0.35) |
|  | T2 Native Mapping, ms | 46.5 (44.4, 51.0) | 48.0 (44.0, 51.4) | 0.97 (0.83-1.13; p=0.70) |
| **Cardiac Fibrosis** | LGE | 0 | 0 | -- |
| **Possible Pericardial Inflammation** | Pericardial Effusion | 6 (24%) | 5 (22%) | 0.47 (0.08-2.71; p=0.41) |

### Palpitations are Not Explained by Arrhythmias on Ambulatory Rhythm Monitoring

Of those included, 38 participants wore and returned an ambulatory rhythm monitor. Lower maximum heart rate, age-predicted maximum heart rate, and adjusted heart rate reserve were all associated with symptoms consistent with our CPET findings (Table 4). One symptomatic individual had a single episode of non-sustained ventricular tachycardia without recorded symptoms or button push; no other clinically significant arrhythmias including atrial fibrillation or atrial flutter were present in either group (Supplemental Table 4). The burden of sinus tachycardia and supraventricular tachycardias was not significantly increased among those with symptoms. Premature ventricular contractions were associated with symptoms, and we could not exclude an association between premature atrial contractions and symptoms (Table 4).

**Table 4:** Ambulatory Rhythm Monitoring Findings by Symptoms (n=38) **Table 4 Legend.** Values are reported as mean±SD or median (interquartile range) for non-normally distributed variables assessed by histogram. Those with self-reported symptoms pressed the symptom button on the monitor more 3.2 times more often (95%CI 2.1-4.7; p<0.001). **Bolded results are p<0.05.**

| Parameter | Symptoms (n=24) | No Symptoms (n=14) | Adjusted OR for symptoms (95%CI; p value) | Adjusted OR for palpitations (95%CI; p value) |
| --- | --- | --- | --- | --- |
| Monitoring Time, days | 4 (3, 13) | 8 (3, 13) | -- |  |
| Average Heart Rate, bpm | 77.5 ± 11.4 | 74.4 ± 4.5 | 1.04 (0.96 to 1.14; p=0.34) | 1.11 (0.98-1.28; p=0.11) |
| Minimum HR, bpm | 50.2 ± 12.5 | 44.9 ± 2.8 | 1.13 (0.96-1.33; p=0.13) | **1.38 (1.04-1.82; p=0.02)** |
| Maximum HR, bpm | 142.2 ± 19.0 | 157.4 ± 21.4 | **1.77 per -10 (1.03 to 3.03; p=0.04)** | **2.60 (1.07-6.34; p=0.04)** |
| Maximum HR, % predicted | 85.1 ± 9.1 | 94.9 ± 11.3 | **3.39 per -10 (1.09 to 10.6; p=0.04)** | **9.15 (1.14-72.9; p=0.04)** |
| Adjusted HRR achieved, % | 70.7 ± 19.7 | 89.9 ± 20.1 | **1.77 per -10 (1.09 to 2.89; p=0.02)** | **4.20 (1.34-13.2; p=0.01)** |
| Heart Rate Variability, SDNN | 143.5 (113.3, 185.5) | 155.7 (137.0, 177.0) | 1.00 per -10 (0.87 to 1.16; p=0.97) | **1.91 (1.08-3.60; p=0.03)** |
| PAC, % burden | 0.02 (0.01, 0.10) | 0.01 (0.01, 0.04) | 1.36 per 10-fold increase (0.58 to 3.20; p=0.48) | 1.05 (0.36-3.05; p=0.93) |
| PVC, % burden | 0.01 (0.01, 0.15) | 0.01 (0, 0.01) | **2.74 per 10-fold increase (1.14 to 6.53; p=0.02)** | 2.20 (0.92-5.31; p=0.08) |
| Sinus tachycardia, % burden | 7 (3, 13) | 4 (3, 6) | 1.15 (0.97 to 1.37; p=0.12) | 1.20 (1.00-1.43; p=0.05) |
| Episodes of SVT* per week | 1.2 (0, 3.5) | 0 (0, 3) | 1.08 (0.91 to 1.28; p=0.39) | 1.15 (0.91-1.46; p=0.25) |
| Episodes of Nonsustained VT | 0 | 1 | -- |  |
| Button Pushes | 2.5 (0.5,7) | 1 (0, 2) | 1.33 (1.00-1.76; p=0.05) | 1.13 (0.93-1.36; p=0.22) |

Symptomatic individuals pressed the button 3.2 times more often (95%CI 2.1-4.7; p<0.001). Button pushes were mostly during sinus rhythm, sinus tachycardia, or supraventricular ectopy (Supplemental Figure). Results were similar considering only those with palpitations.

### Ambulatory Rhythm Monitoring Correlates of Chronotropic Incompetence on CPET

CPET peak HR correlated with maximum sinus HR during ambulatory monitoring (Pearson’s r=0.71; p<0.001), with ambulatory peak HR 29 bpm lower among those with chronotropic incompetence (95%CI 13-45; p<0.001). Chronotropic incompetence was associated with 12.6 bpm higher minimum HR (95%CI 3-22; p=0.01) and 59ms lower HR variability by standard deviation n-to-n (95%CI 24-95; p=0.002; Supplemental Table 4). PR intervals were not significantly longer among those with chronotropic incompetence (171ms vs 168ms; p=0.72). One symptomatic individual had 2^nd^ degree Mobitz type 1 (normal finding), and no participants had 2^nd^ degree Mobitz type 2 or 3^rd^ degree heart block.

### Markers of Inflammation Early in PASC are Associated with Exercise Capacity and Pericardial Effusions More than 1 Year Later

Markers of inflammation in the blood (hsCRP, IL-6, TNF) and SARS-CoV-2 RBD IgG level, but not hs-troponin or NT-pro-BNP measured at 3-9 months after infection are negatively correlated with peak VO_2_ more than one year after infection (Figure 3). After adjustment, peak VO_2_ was 6.2 ml/kg/min lower per doubling of TNF (95%CI 0.6-11.8; p=0.03) and 1.8 ml/kg/min lower per doubling of hsCRP (95%CI 0.8-2.9; p=0.001).

**Figure 3.**
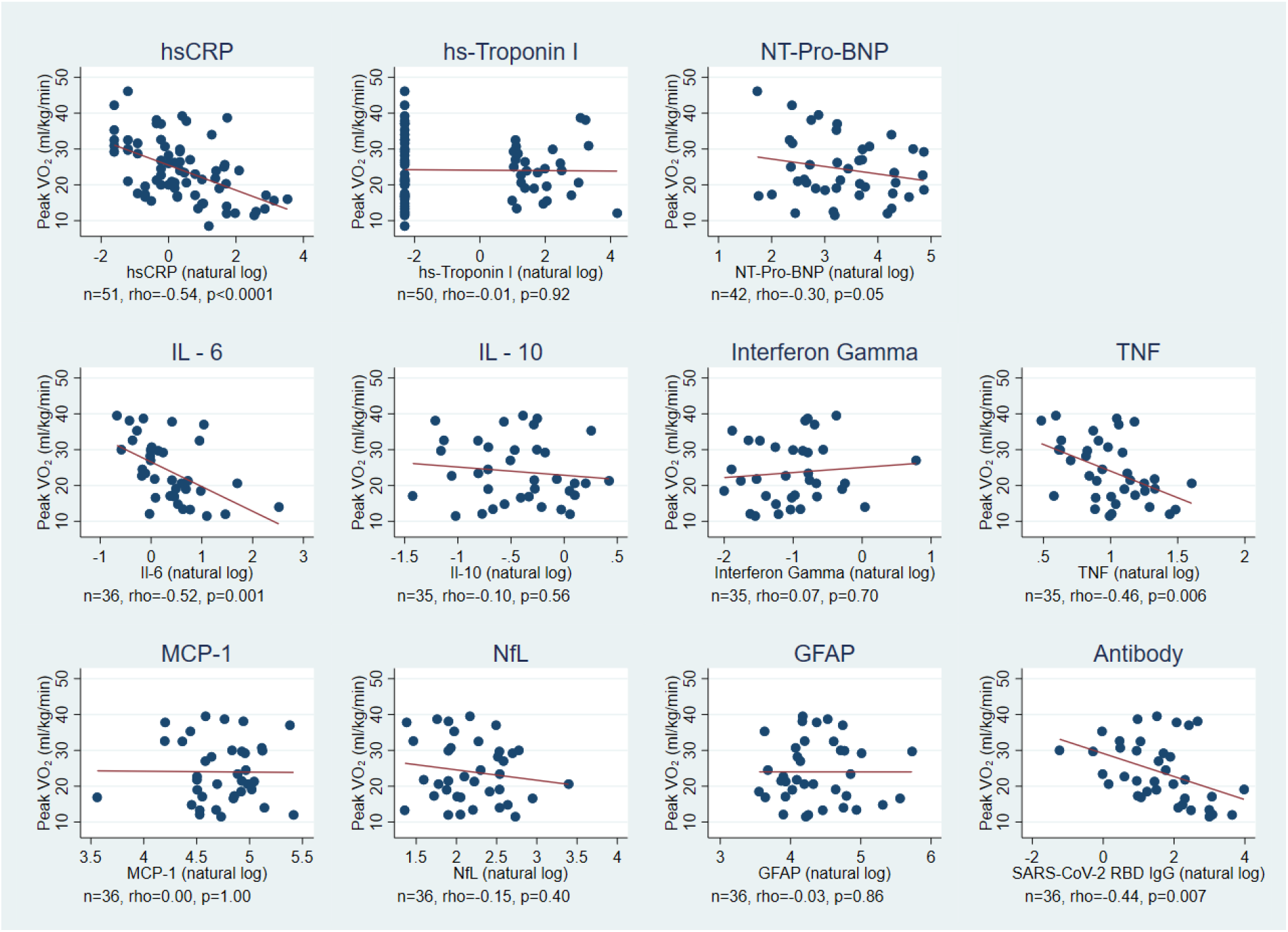
Correlations between Peak VO_2_ and Previously Measured Biomarkers. Figure 3 **Legend.** Scatterplots and linear trend lines of peak VO_2_ (measured at ∼18 months) by natural log of previously measured biomarker levels with unadjusted Pearson’s rho correlations and p-values listed (top row, median 6 months after SARS-CoV-2 infection; bottom two rows median 3.5 months after SARS-CoV-2 infection). Prior hsCRP, IL-6, TNF and SARS-CoV-2 receptor binding domain antibody levels were correlated with subsequent peak VO_2_. All antibody levels were measured prior to vaccination.

Longitudinal serum biomarkers of inflammation, neurologic injury, and SARS-CoV-2 RBD IgG were measured at <90 days from SARS-CoV-2 acute infection (median 52 days) and between 90-150 days (median 124 days) in 36 participants who underwent CPET (Figure 4), all prior to vaccination. SARS-CoV-2 IgG (2.99-fold higher mean ratio, 95%CI 1.41-6.33; p=0.004) and TNF (1.34-fold higher mean ratio, 95%CI 1.11-1.61; p=0.002) were higher at <90 days among those with reduced exercise capacity. At 90-150 days, only SARS-CoV-2 IgG (2.12-fold higher mean ratio, 95%CI 1.02-4.43; p=0.04) remained statistically significant, although after adjustment it was no longer statistically significant (1.1 ml/kg/min per doubling, 95%CI -0.3 to 2.4; p=0.11). We could not exclude an effect of IL-6 (1.34-fold higher mean ratio, 95%CI 0.92-1.96; p=0.11; adjusted 2.1 ml/kg/min per doubling, 95%CI -0.5-4.6; p=0.11). Except for IL-6, all other biomarkers decreased over time regardless of eventual exercise capacity.

**Figure 4.**
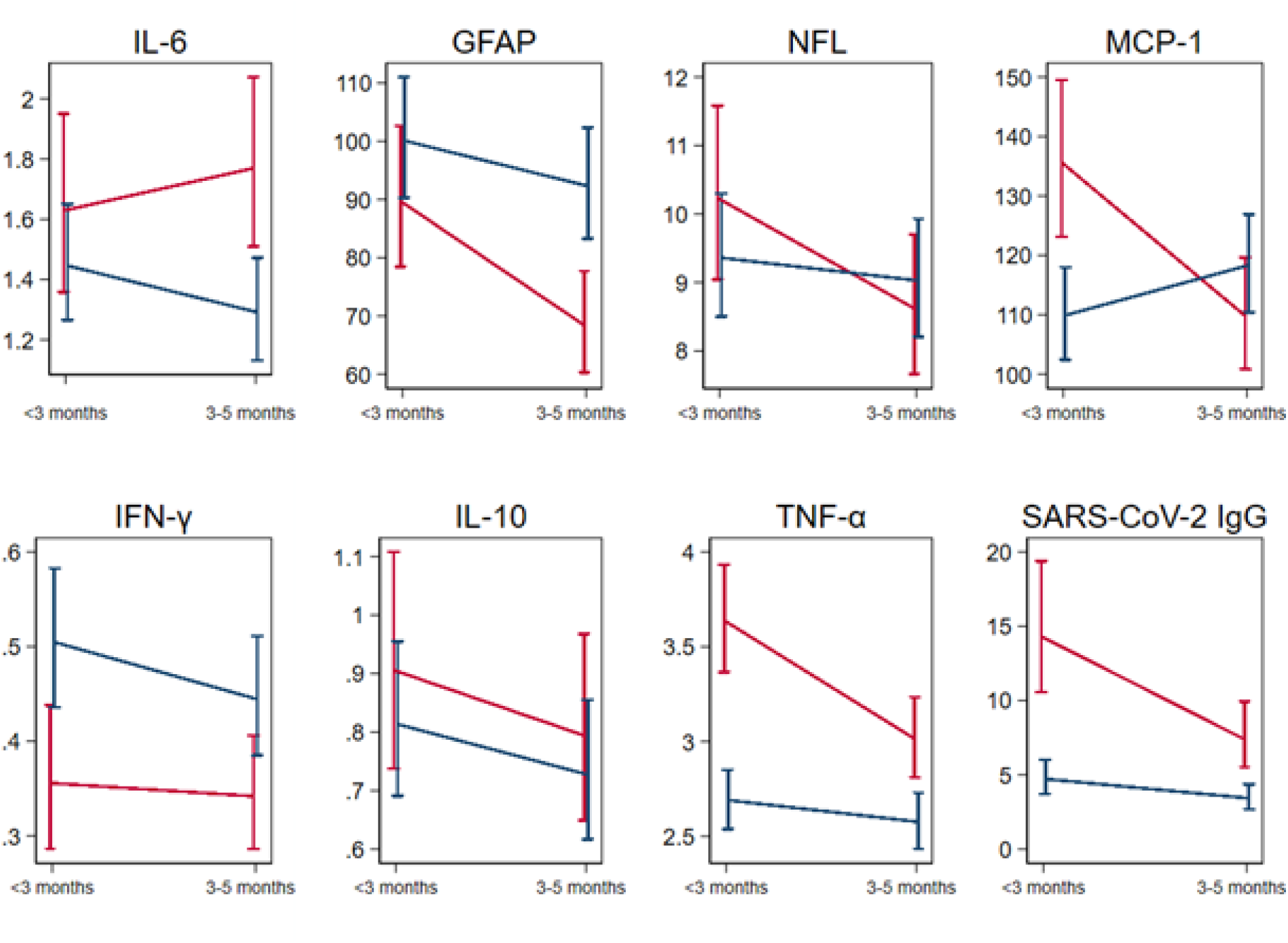
Change in Biomarkers Early Post-Infection by Eventual Exercise Capacity at 18 months (n=35) Figure 4 **Legend:** Mean ± standard error of the mean for serum biomarkers over time among those with reduced exercise capacity (red) and preserved exercise capacity (blue) measured at <90 days from SARS-CoV-2 acute infection (median 52 days from symptom onset) and between 90-150 days (median 124 days from symptom onset) in 35 participants who underwent CPET. Inflammatory markers decreased over time, except for IL-6 among those with reduced exercise capacity. MCP-1, TNF, and IgG were higher early among those with reduced exercise capacity. TNF and IgG remained higher and GFAP became lower among those with reduced exercise capacity at the second time point.

## Discussion

We demonstrate that clinically meaningful reductions in objective exercise capacity are associated with LC symptoms more than 1 year after SARS-CoV-2 infection. Our findings suggest that chronotropic incompetence contributes to exercise limitations in LC. We found elevated inflammatory markers and SARS-CoV-2 antibody levels early in PASC are associated with reduced exercise capacity more than a year later. We did not find evidence of myocarditis, cardiac dysfunction, or clinically significant arrhythmias. Finally, our study validates that CPET allows objective measurement of patient-reported exercise intolerance and therefore may be useful for interventional trials of therapeutics for LC.

### Connections between Inflammation, Reduced Exercise Capacity, and Autonomic Responses

Our study extends prior findings that inflammatory markers including hsCRP, IL-6, and TNF are negatively correlated with peak VO_2_ early after COVID-19 hospitalization [17] to >1 year after infection and those not hospitalized. This correlation may reflect a common cause for inflammation and exercise limitations in PASC (i.e., viral persistence [18], immune activation [19]), or these markers could be on the causal path from infection to symptoms and reduced exercise capacity. Endothelial and coronary microvascular dysfunction occur in PASC [20–23] and are associated with chronotropic incompetence and inflammation [24, 25]. Inflammation may alter autonomic function which could explain reduced exercise capacity and chronotropic incompetence.

Data from animal models support the hypothesis that chronotropy may be related to inflammatory signals and endothelial functions. Apart from COVID-19, IL-6 impairs chronotropic responses to autonomic signaling in mice [26] and may regulate energy allocation during exercise [27]. In addition, IL-6 and TNF impair endothelial function in animal models via increasing oxidative stress and suppressing endothelial nitric oxide synthase pathways [28].

Ideally, clinical trials should evaluate whether anti-inflammatory strategies improve chronotropy and endothelial function, as both impact exercise capacity and risk of cardiovascular disease.

### Autonomic Function, Sinus Node Function, and Inflammation in PASC

Altered autonomic function is a possible unifying explanation for our and others’ CPET findings in PASC including altered peripheral oxygen extraction [29], preload failure [30–32], and disordered breathing [30]. Orthostatic intolerance, an autonomic symptom, occurs in PASC [33], and skin biopsies suggest small fiber neuropathy in LC-associated postural-orthostatic tachycardia syndrome [34, 35]. Effects on brainstem regulatory regions or the amygdala [36] could also modify autonomic responses to exercise.

An alternative hypothesis is that SARS-CoV-2 could alter sinus node function. Autopsy studied have not specifically examined sinus node tissue for evidence of persistent viral infection of the sinus node [37], but hamster models suggest that SARS-CoV-2 can infect hamster sinoatrial node cells and *in vitro* sinoatrial-like pacemaker cells resulting in altered calcium handling, activated inflammatory pathways, and induced ferroptosis [38]. Sinus node remodeling may reduce sinus node reserve in heart failure [39]. Although we did not find evidence of cardiac fibrosis or sinus node dysfunction, we cannot fully exclude that sinus node dysfunction may contribute to CI. A combination of autopsy studies that specifically examine sinoatrial tissue and clinical studies may be necessary to identify the relevant mechanisms.

Inflammation modifies autonomic and chronotropic responses. Young adults recovering from SARS-CoV-2 have elevated sympathetic activation at rest [40]. Chronic inflammation in other settings is associated with parasympathetic and sympathetic imbalance, chronotropic incompetence and reduced exercise capacity [24, 41]. Thus, chronic inflammation could blunt chronotropy in PASC even without autonomic nervous system or sinus node damage.

### Other Studies of PASC using CPET

Our study is consistent with others that have reported lower peak VO_2_ among those with PASC compared to recovered individuals mostly at 3-6 months after severe COVID-19, which we summarized in a systematic review and meta-analysis [10]. Our findings build upon earlier studies by (1) demonstrating reduced peak VO_2_ and chronotropic incompetence much later after infection (2) including evaluation of cardiac inflammation, structural heart disease and arrhythmias, (3) adjusting for confounders, (4) including recovered persons as comparators, and (5) demonstrating associations with longitudinal biomarkers.

Differences in classification of exercise limitations across CPET studies of PASC may arise from selection bias, confounding, different CPET protocols, and different interpretation algorithms. Deconditioning, which contributes to reduced exercise capacity after any illness, may be misidentified from noninvasive CPET, and has been commonly reported 3-6 months after hospitalization [10]. Although reductions in physical activity after COVID-19 [42] suggest deconditioning contributes, our findings argue against deconditioning as the only explanation for most individuals as deconditioning more commonly demonstrates an accelerated rather than a blunted heart rate response.

Five other studies have also found that chronotropic incompetence contributes to exercise limitations in PASC [32, 42–45]. Chronotropic incompetence may be underestimated in some studies as sensitivity and specificity vary with exercise modality and protocol and including sub- maximal tests or patients on beta-blockers reduces specificity. Diagnosing chronotropic incompetence may have prognostic implications: it is associated with incident cardiovascular disease, sudden death, and all-cause mortality among men without coronary artery disease [46–48].

Impaired peripheral oxygen extraction, best assessed with invasive CPET, may also contribute to exercise limitations in PASC [29], perhaps via changes in autonomic regulation of microcirculatory function [49] or altered metabolism [50]. We did not find differences in VO_2_/work slope, a noninvasive correlate of measured oxygen extraction. Although not observed among our participants, dysfunctional (rapid, erratic) breathing and exercise hyperventilation may contribute to dyspnea in PASC [30, 51, 52].

### CMR and Ambulatory Rhythm Monitoring Findings

CMR findings suggestive of myocarditis without cardiac dysfunction may be present in the early post-acute period [5, 8, 53]. Consistent with studies at later time points [6, 7], we did not find evidence of abnormal function or LGE, suggesting that myocarditis is unlikely to explain symptoms in most with PASC.

Our findings are consistent with two studies that did not find arrhythmias in early PASC [54, 55]. In contrast to a study in early PASC [56], inappropriate sinus tachycardia was present only in one individual (without symptoms). Therefore, arrhythmias and inappropriate sinus tachycardia are unlikely to explain symptoms among most individuals with PASC.

### Implications for Therapy

Investigation into mechanisms of PASC may benefit from proof-of-concept approaches to identify potential therapies. Although vaccination reduces the risk of PASC [57, 58] and the newer circulating variants may be associated with lower risk of Long COVID [59], there is no data regarding whether anti-viral, anti-inflammatory, or anti-coagulant strategies improve exercise capacity in PASC. In chronotropic incompetence separate from COVID-19, chronic supervised exercise is the only intervention demonstrated to improve exercise capacity and surrogates of autonomic function [60–62]. Exercise is an effective treatment for postural orthostatic tachycardia syndrome, which may also be related to post-viral alterations in autonomic responses to stress and occurs in PASC [63, 64]. Reports of post-exertional malaise or symptom exacerbation (PEM/PESE) in PASC overlapping with ME/CFS [65–67] mean exercise-based interventions should be considered with caution. A study of a six-week structured pacing intervention improved physical activity levels and reduced PESE [68], and another study found that supervised exercise may be helpful rather than harmful in PASC [69].

### Limitations

The main limitations of this observational study arise from the small sample size, non- probabilistic sampling, and cross-sectional cardiac measures. Secondly, the difference in peak VO_2_ was sensitive to the case definition, but our definition is consistent with current consensus definitions [12]. Volunteer bias may result in overestimated prevalence and magnitude of reduced exercise capacity but should not affect classification of limitations. We did not include an uninfected comparator group and nearly all individuals were unvaccinated at the time of initial SARS-CoV-2 infection. Although we excluded those with cardiac disease, adjusted for measured confounders, and conducted sensitivity analyses adjusting for additional confounders, unmeasured residual confounders including pre-COVID fitness remain. Adjustment (in BMI, for example) may not have fully accounted for confounding. Misclassification of exercise limitations could occur since we did not perform invasive CPET, stress echocardiography, stress CMR, or stress ventriculography. Lastly, we lacked contemporaneous biomarker data with CPETs to ascertain whether a transient inflammatory process or ongoing inflammation is more likely.

### Conclusions

In conclusion, more than 1 year after pre-vaccine index SARS-CoV-2 infection, reduced exercise capacity on CPET is associated with LC symptoms, chronotropic incompetence, and higher inflammatory markers and antibody levels in the early post-acute period, but not evidence of myocarditis or arrhythmias. Further investigation into mechanisms of cardiopulmonary PASC should include evaluation of inflammatory pathways, chronotropic function, and the autonomic nervous system to identify therapeutic targets.

## Data Availability

All data produced in the present study are available upon reasonable request to the authors.

## Acknowledgements

We would like to thank the research participants and the members of the LIINC study team. We would like to thank Dr. Kara Lynch and Dr. Alan Wu for their assistance with measuring cardiac biomarkers and hsCRP. We would also like to acknowledge support from Jeremy Lambert from Quanterix (Billerica, MA) and Patrick Kaiser from BardyDx, a division of Hillrom (Bellevue, Washington). We acknowledge the contributions of the UCSF Clinical and Translational Science Unit. This study was funded by philanthropic gifts from Charles W. Swanson and the Ed and Pearl Fein Foundation, research grants from the NIH/NLBI including L30 HL159695 and K12 HL143961, and internal funds from the Division of Cardiology at Zuckerberg San Francisco General. This work was assisted in part by a CFAR-ARI Boost Award from the UCSF AIDS Research Institute. MSD is supported by K12HL143961. MJP is supported by K23AI157875. JDK is supported by NIH/NIAID K23AI135037. TJH is supported by NIH/NIAID 3R01A1141003-03S1. PYH is supported by NIH/NAID 2K24AI112393-06. This publication was supported by the National Center for Advancing Translational Sciences, National Institutes of Health, through UCSF-CTSI Grant Number UL1TR001872. Its contents are solely the responsibility of the authors and do not necessarily represent the official views of the NIH. The funders had no role in study design, data collection and analysis, decision to publish, or preparation of the manuscript.

## Author Contributions

MSD designed the study, obtained IRB approval, acquired data, analyzed data, interpreted the findings, and wrote the first draft of the manuscript. MJP assisted in study design, participant recruitment, data interpretation, and critical revision of the manuscript. PK and YJL conducted CMR measurements, drafted the MRI methods, and contributed to interpretation of the MRI findings. CH led the ambulatory rhythm monitoring measurements and participated in interpretation of the findings. DL and ES participated in the cardiopulmonary exercise testing measurements, drafting of the CPET methods, CPET interpretation, and provided input on the manuscript. SS and VMA participated in measurements, data management, and revisions of the manuscript. DVG participated in statistical design, analysis and visualization. SL, SAG, and RH participated in participant recruitment, measurement, and data management. AC, BCY, JWW, and CJP participated in biomarker measurement and provided input on drafts of the manuscript. JDK, TJH, and JNM participated in the study design, interpretation of results, and revision of the manuscript. CSL and DJG provided funding and participated in designing the study, interpreting results, and revising the manuscript. SGD and PYH provided funding, oversight, and mentorship for the first two authors and participated in study design, interpretation of results, and critical manuscript revision.

## Competing Interests statement

AC, BCY, JWW, and CJP are employees of Monogram Biosciences, Inc., a division of LabCorp. PYH has received modest honoraria from Gilead and Merck and research grant from Novartis unrelated to the submitted work. All other authors report no other competing interests.

## Notes

### Author Declarations

IRB of University of California, San Francisco gave ethical approval for this work.

### Summary of Updates

Updated with complete CPET cohort and updated figures and tables.

