## Supplemental Materials for "Reduced exercise capacity, chronotropic incompetence, and early systemic inflammation in cardiopulmonary phenotype Long COVID"

**SUPPLEMENTAL MATERIALS**

**Supplemental Methods:**

*Biomarkers*

Samples were batch processed for measurement of high-sensitivity troponin I (hs-troponin; ADVIA Centaur® High-Sensitivity Troponin I (TNIH) assay), N-terminal prohormone b-type natriuretic protein (NT-pro-BNP; Roche Cobas 6000 Elecsys® proBNP II assay), and high-sensitivity c-reactive protein (hsCRP; ADVIA® Chemistry CardioPhase™ High Sensitivity C-Reactive Protein assay). Most participants had antibodies and additional markers measured at samples from two earlier time points (<90 days and 90-150 days after infection) including IL-6, interleukin 10 (IL-10), glial fibrillary acidic protein (GFAP), neurofilament light chain (NfL), monocyte chemoattractant protein-1 (MCP-1), interferon gamma (IFN -γ), and tumor necrosis factor alpha (TNF-α), and SARS-CoV-2 receptor-binding domain (RBD) immunoglobulin G (IgG) (18). These samples were assayed by Monogram Biosciences (South San Francisco, CA) using the Quanterix Simoa® platform with Simoa® Assay Kits from Quanterix Corporation (Billerica, MA) blinded with respect to patient and clinical information, and assay performance was consistent with manufacturers' specifications.

Hematocrits were collected immediately before CMR with patient sitting right outside the MRI suite.

*Cardiac MRI imaging sequences:*

The protocol consisted of acquisition of the following sequences after multiplane localizers prior to gadolinium injection: fast imaging employing steady state acquisition (FIESTA) cine in the axial short axis planes, pre-contrast T1 mapping sequences using the MOLLI 5-(3)-3 technique, as well as pre-contrast T2 mapping at the basal, mid, and apical short axis planes and T2 fat-saturated weighted black blood spin-echo images in the short axis plane. 8-10 minutes after intravenous gadolinium injection, phase sensitive inversion recovery (PSIR) late gadolinium enhancement imaging in short axis full stack, 4 chamber full stack, three slices of 2 chamber images, and post-contrast T1 mapping sequences were obtained. Inversion times were individualized to null the myocardium.

Arterys software was used for T1 and T2 mapping and ECV calculation using pre- and post-contrast MOLLI sequences.

*Cardiopulmonary Exercise Testing*

First, baseline ECG, blood pressure, and rest spirometry including maximum voluntary ventilation (MVV) were measured. We determined the work increase per 1 minute step based on the expected peak $VO_2$ from the MVV for a goal 10 minute test, rounded to 5 Watts/min increments based on reported exercise (range 10-30W/min) in accordance with guidelines.(58) Participants underwent a 2-minute rest phase, 2-minute no resistance warm up, and then 1-minute steps. Breath-by-breath oxygen consumption ($VO_2$) and carbon dioxide production

($VCO_2$) were measured continuously. Participants were blinded to time, wattage, and peak $VO_2$ during the test and encouraged to maintain a cadence of ~60 cycles per minute and exercise to their maximum ability, with the test stopped prematurely for severe hypertension, relative hypotension, moderate to severe angina, ventricular tachycardia or couplets, or ischemic changes. Reason for stopping and exertional symptoms were recorded. Exercise effort was assessed by Borg Scale and respiratory exchange ratio (RER). Anaerobic threshold was determined manually by the exercise physiologist using the slope method.

*Classification of CPET Limitations*

We considered ventilatory limitation if rest spirometry was abnormal, tidal volume did not double over baseline reaching at least 50% of FVC, breathing reserve was <30%, dead space ratio ($V_D/V_T$) was >0.2, or desaturation occurred. We classified participants as having a cardiac limitation if there were ischemic ECG changes, the oxygen pulse was reduced and plateaued early, AT occurred at less than 40% of predicted maximal $VO_2$, and if ventilatory efficiency slope ($VE/VCO_2$) was <32. We considered exercise limitation to be likely due to deconditioning, obesity or impaired gas exchange if exercise capacity was reduced without cardiac or ventilatory limitation.

**Supplemental Table 1: Symptom pattern by individual symptoms**

| | Prior to COVID (excluded) | Never reported | Reported at 1st visit but not 2nd visit | Reported at 2nd visit but not first visit | Reported at both visits |
|---|---|---|---|---|---|
| Any symptom (including fatigue) | n/a | 15 | 7 | 5 | 33 |
| Chest pain, dyspnea, or palpitations | n/a | 20 | 9 | 8 | 23 |
| Chest pain | 1 | 41 | 3 | 8 | 7 |
| Dyspnea | 1 | 30 | 9 | 3 | 17 |
| Palpitations | 4 | 32 | 5 | 8 | 11 |
| Fatigue | 2 | 20 | 7 | 11 | 20 |

**Supplemental Table 1 Legend:** Trajectory of Symptoms by specific symptoms and whether they were present prior to COVID-19, never reported, reported at first visit and resolved prior to second visit, developed after the first visit, or were persistent. Note that those with symptoms prior to COVID are excluded for individual symptoms.

**Supplemental Table 2. Additional CPET parameters including resting echocardiogram and spirometry**

| | Symptoms (n=37) | No Symptoms (n=19) | OR (95%CI; p value) |
|---|---|---|---|
| Additional CPET Parameters | | | |
| $VO_2AT$, ml/kg/min | 10.9 (9.4, 13.1) | 13.6 (11.5, 17.4) | 0.94 (0.81 to 1.10; p=0.49) |
| $VO_2AT$ % Predicted $pVO_2$ | 56.5 ± 11.2 | 50.8 ± 10.0 | 1.05 (0.97-1.13; p=0.21) |
| Weber Class<br>A ($pVO_2$>20)<br>B ($pVO_2$ 16-20)<br>C ($pVO_2$ 10-16) | <br>22 (61%)<br>8 (22%)<br>7 (19%) | <br>17 (89%)<br>1 (5%)<br>1 (5%) | p=0.16<br>Ref<br>7.2 (0.6-89)<br>6.5 (0.41-103) |
| Rest Respiratory Rate | 17.2 ± 4.7 | 14.6 ± 6.5 | 1.14 (0.99-1.31; p =0.07 |
| Tidal Volume at Peak (% FVC) | 51.7±9.1 | 57.1±12.1 | 0.95 (0.88-1.03; p=0.19) |
| Dead Space at Peak (% $V_T$) | 17.6±3.7 | 16.1±2.9 | 1.27 (0.97-1.68; p=0.08) |
| Vent. Class[a]<br>I<br>II | <br>23 (72%)<br>9 (28%) | <br>12 (75%)<br>4 (25%) | <br><br>1.55 (0.31-7.72; p=0.59) |
| **Rate Pressure Product, mmHg*bpm** | **25469± 7023** | **29712± 4088** | **1.29 per -1000 (1.01 to 1.40; p=0.04)** |
| Rest Echocardiographic Parameters | | | |
| Left Ventricular Ejection Fraction, % | 64.9±5.4 | 65.3±5.0 | 0.96 (0.84 to 1.08; p=0.49) |
| LV Diastolic Function<br>Indeterminate<br>Mild | <br>6 (17%)<br>5 (14%) | <br>1 (5%)<br>1 (5%) | p=0.08<br><br>6.9 (0.62-75.8) |
| LV Strain, % | -20.0±2.6% | -19.7±1.9% | 1.01 (0.77 to 1.32; p=0.96) |
| RV Strain, % | -24.6±5.5% | -23.9±3.3% | 0.99 (0.85-1.15; p=0.90) |
| TAPSE, cm | 2.29±0.38 | 2.28±0.23 | 1.01 per -10mm (0.85-1.21; p=0.91) |
| Pulmonary Artery Pressure, mm Hg | 24.3±4.6 | 21.6±2.0 | 1.36 (1.02 to 1.81; p=0.04) |
| Pericardial Effusions | 3 (8%) | 0 | n/a |
| Rest Spirometry | | | |
| Forced Vital Capacity, L | 3.8±1.0 | 4.1±0.6 | 0.82 (0.32-2.07; p=0.67) |
| Forced Vital Capacity, % Pred | 97±17 | 101±12 | 0.97 (0.93-1.02; p=0.26) |
| Forced Expiratory Volume in 1 second, % Pred | 102±18 | 100±12 | 1.01 (0.97-1.05; p=0.64) |
| $FEV_1$/FVC, % Pred | 104±10 | 101±5 | 1.12 (1.01-1.25; p=0.03) |
| Slow Vital Capacity, % Pred | 100±16 | 104±16 | 0.99 (0.95-1.03; p=0.55) |
| Inspiratory Capacity, % Pred | 104±27 | 110±25 | 0.98 (0.95-1.01; p=0.18) |
| Maximum Voluntary Ventilation (measured), L/min | 130±39 | 139±28 | 1.00 (0.98-1.03; p=0.90) |

| | | | |
|---|---|---|---|
| Maximum Voluntary Ventilation, % Pred | 105±25 | 104±17 | 1.00 (0.97-1.03; p=0.96) |

**Supplemental Table 2 Legend:** Echocardiographic parameters by cardiopulmonary symptom status at echo visit (median 6 months). Spirometry was performed at the time of CPET (median 18 months). Odds ratios are adjusted for age, sex, time since SARS-CoV-2 infection, BMI, hospitalization for COVID-19; no change in sensitivity analysis additionally adjusting for medical history. Mild diastolic dysfunction and indeterminate were combined due to small numbers and non-convergence of the model. Abbreviations: AT=Anaerobic threshold; bpm=beats per minute; FVC=Forced Vital Capacity; HR=heart rate; DBP=diastolic blood pressure; MVV=maximal voluntary ventilation; SBP=systolic blood pressure; $V_D/V_T$=Dead space ratio; $V_E$ = minute ventilation; $VCO_2$=carbon dioxide production; $pVO_2$=peak oxygen consumption ($VO_2$); Vent=Ventilatory

**Supplemental Table 3. Differences in Peak $VO_2$ by Different Means of Classifying Symptoms**

| | | Symptom Present | Symptom Absent | Adjusted Difference |
|---|---|---|---|---|
| Dyspnea, Chest Pain, Palpitations, or Fatigue at Visit 2 (n=27) | **$VO_2$ (ml/kg/min)** | **22.7±8.1** | **29.6±7.0** | **-5.2 (-8.3 to -2.1; p=0.001)** |
| | **$VO_2$, % predicted** | **92 ± 22** | **107 ± 22** | **-17 (-30 to -4; p=0.01)** |
| Dyspnea, Chest Pain, or Palpitations at Visit 2 (n=30) | $VO_2$ (ml/kg/min) | 23.3 ± 8.7 | 27.0 ± 7.8 | -2.8 (-6.2 to 0.6; p=0.10) |
| | $VO_2$, % predicted | 95 ± 23 | 100 ± 23 | -6 (-20 to 7, p=0.36) |
| Self-Reported Reduced Exercise Capacity at Visit 2 (n=31) | $VO_2$ (ml/kg/min) | 24.7 ± 8.6 | 25.5 ± 8.3 | -0.9 (-4.1 to 2.3; p =0.59) |
| | $VO_2$, % predicted | 94 ± 23 | 101 ± 23 | -7.9 (-20 to 5; p=0.21) |
| Persistent (n=32) vs Never Symptoms (n=13) | **$VO_2$ (ml/kg/min)** | **22.7 ± 8.5** | **29.4 ± 8.4** | **-5.9 (-9.8 to -2.0; p=0.004)** |
| | $VO_2$, % predicted | 94 ± 23 | 110 ± 23 | -16 (-32 to 0.4; p=0.06) |
| Symptoms at Visit 1, median 6 months (n= 31) | $VO_2$ (ml/kg/min) | 23.0 ± 8.4 | 27.6 ± 7.9 | -2.9 (-6.3 to 0.5; p=0.10) |
| | $VO_2$, % predicted | 95 ± 23 | 100 ± 23 | -3.3 (-17 to 10; p=0.63) |

**Supplemental Table 3 Legend:** Sensitivity analysis of peak $VO_2$ (ml/kg/min and % predicted) using different symptom definitions to classify participants. N listed for the number with that symptom finding.

**Supplemental Table 4: Heart Rate Parameters by Chronotropy and Exercise Capacity**

| Measure | | Value, mean±SD | Adjusted Difference (95%CI; p value) |
|---|---|---|---|
| CPET Peak HR During Exercise, bpm | CI | 119±14 | -43 (-50 to -36; p<0.001) |
| | Reduced HR | 135±12 | -29 (-31 to -24; p<0.001) |
| | Normal | 170±13 | |
| CPET Peak HR, %Age Predicted Max | CI | 73±5 | -27 (-32 to -21; p<0.001) |
| | Reduced HR | 84±3 | -17 (-20 to -14; p<0.001) |
| | Normal | 100±5 | |
| CPET AHRR, % | CI | 48±7 | -49 (-56 to -42; p<0.001) |
| | Reduced HR | 65±10 | -34 (-41 to -27; p<0.001) |
| | Normal | 99±10 | |
| CPET HR Recovery at 1 minute, bpm | CI | 9.1±6.1 | -7.9 (-15.3 to -0.4; p=0.04) |
| | Reduced HR | 10.9±8.3 | -4.5 (-11.6 to 2.7; p=0.21) |
| | Normal | 17.1±8.5 | |
| Ambulatory Max sinus HR, bpm | CI | 129±20 | -25 (-39 to -11; p=0.002) |
| | Reduced HR | 145±19 | -4.8 (-17 to 8; p=0.43) |
| | Normal | 158±17 | |
| Ambulatory Max HR, % Age Predicted Max | CI | 81±12 | -10.3 (-19.7 to -0.9; p=0.04) |
| | Reduced HR | 87±14 | -2.0 (-13.0 to 9.0; p=0.68) |
| | Normal | 95±7 | |
| Ambulatory AHRR*, % | CI | 57±22 | -31 (-46 to -16; p=0.001) |
| | Reduced HR | 79±25 | -6.0 (-20 to 8.1; p=0.38) |
| | Normal | 90±13 | |
| Ambulatory Minimum HR, bpm | CI | 57±16 | 11.5 (2.4 to 20.6; p=0.02) |
| | Reduced HR | 46±6 | 1.0 (-4.1 to 6.2; p = 0.68) |
| | Normal | 44±5 | |
| Ambulatory Average HR, bpm | CI | 81±13 | 7.8 (0.2 to 15.4; p=0.05) |
| | Reduced HR | 76±11 | 2.5 (-6.4 to 10.9; p=0.59) |
| | Normal | 73±6 | |
| Ambulatory HR Variability SDNN | CI | 107±37 | -62 (-98 to -25; p=0.003) |
| | Reduced HR | 178±73 | 0 (-62 to 61; p=0.99) |
| | Normal | 167±41 | |

**Supplemental Table 4 Legend.** The first row of each measure is the mean±SD for those with chronotropic incompetence ("CI", $VO_2$ <85%, AHRR<80%, and no alternative findings, n=12); the second row is the mean±SD for those with a reduced chronotropic response ($VO_2$ ≥85% and AHRR<80%, n=13) and the third row is those with peak $VO_2$ ≥85% and AHRR≥80% (n=23). Those with limited exercise capacity for other reasons were excluded. Adjusted differences are compared to those with normal exercise capacity and heart rate response during exercise. Results were similar whether considering absolute heart rate, percent of age predicted, or adjusted heart rate reserve for both CPET and ambulatory measurements. Ambulatory adjusted heart rate reserve uses (max sinus heart rate – average heart rate)/(220-age-average heart rate). Abbreviations: HR=heart rate, bpm=beats per minute, CI=chronotropic incompetence, AHRR=adjusted heart rate reserve, SDNN=standard deviation n-to-n.

**Supplemental Figure 1. Cardiac Rhythms During Button Pushes (n=38)**

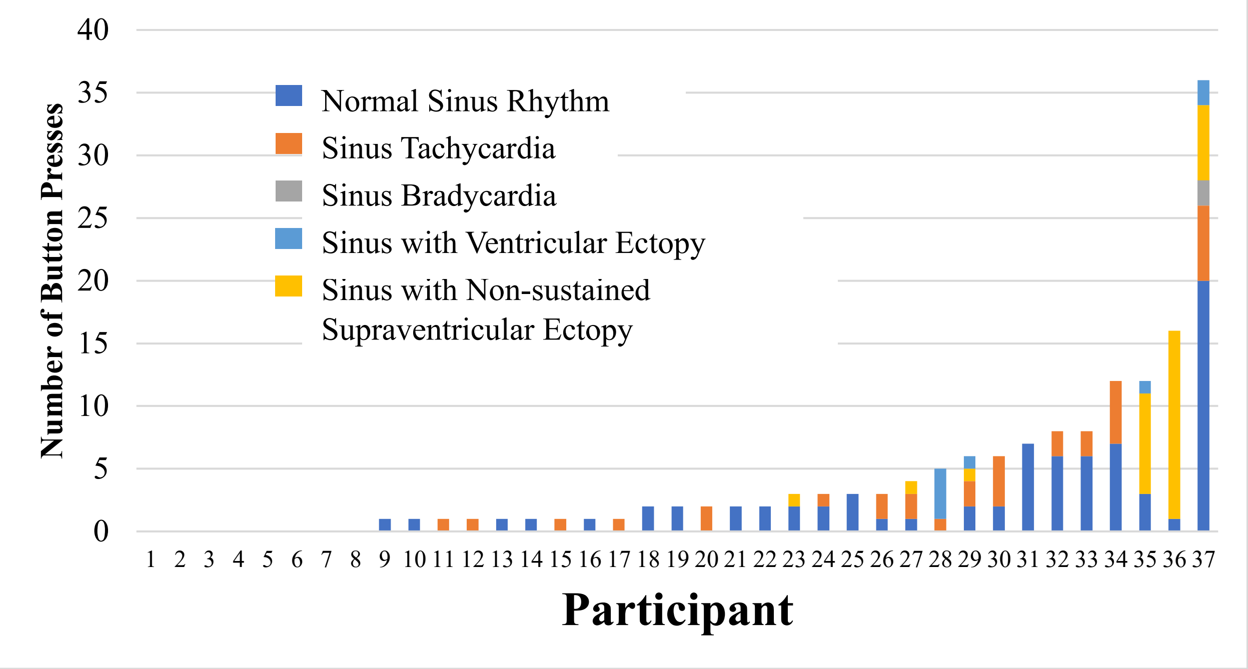


**Supplemental Figure 1 Legend:** Individual participant analysis of number of button pushes and assocaited rhythm with each participant on the x axis and the number of times that individual pushed the symptom button on the y axis with each identified rhythm coded by color. Most button pushes were associated with sinus rhythm or sinus tachycardia, with supraventricular ectopy (premature atrial contractions) present among a few individuals especially the 3 with the most button pushes.
